# A fourth dose of the mRNA-1273 SARS-CoV-2 vaccine improves serum neutralization against the delta variant in kidney transplant recipients

**DOI:** 10.1101/2021.11.25.21266704

**Authors:** Ilies Benotmane, Timothée Bruel, Delphine Planas, Samira Fafi-Kremer, Olivier Schwartz, Sophie Caillard

## Abstract

In immunocompetent subjects, the effectiveness of SARS-CoV-2 vaccines against the delta variant appears three- to five-fold lower than that observed against the alpha variant. Additionally, three doses of SARS-CoV-2 mRNA-based vaccines might be unable to elicit a sufficient immune response against any variant in immunocompromised kidney transplant recipients. This study describes the kinetics of the neutralizing antibody (NAbs) response against the delta strain before and after a fourth dose of a mRNA vaccine in 67 kidney transplant recipients who had experienced a weak antibody response after three doses. While only 16% of patients harbored NAbs against the delta strain prior to the fourth injection – this percentage raised to 66% afterwards. We also found that, after the fourth dose, the NAbs titer increased significantly (p=0.0001) from <7.5 (IQR : <7.5−15.1) to 47.1 (IQR <7.5−284.2). Collectively, our data indicate that a fourth dose of the mRNA-1273 vaccine in kidney transplant recipients with a weak antibody response after three previous doses improves serum neutralization against the delta variant.

---

Research has suggested that even three doses of SARS-CoV-2 mRNA-based vaccines might be unable to elicit a sufficient immune response in immunocompromised kidney transplant recipients.^1–3^ As a result, in June 2021 the French health authorities allowed offering a fourth vaccine dose to weak responder solid organ transplant recipients.While the antibody response mounted by the Pfizer and AstraZeneca vaccines in immunocompetent subjects seems sufficient to neutralize the currently dominant SARS-CoV-2 strain (delta variant),^4^ the effectiveness appears three- to five-fold lower than that observed against the alpha variant.^5,6^ In addition, standard vaccination schemes are beset by low immunogenicity in immunocompromised subjects – who remain prone to develop severe COVID-19.^4^ The purpose of this study is to describe the kinetics of the neutralizing antibody response against the delta strain before and after a fourth dose of the mRNA-1273 (Moderna) vaccine in kidney transplant recipients who had experienced a weak antibody response after three previous doses. We also assessed the correlation between this neutralizing activity and levels of IgG against the receptor binding domain (RBD) of the spike (S) protein.

## Results

Sixty-seven kidney transplant recipients (median age: 56.6 years; interquartile range [IQR]: 47−64.6 years; 61.2% men) showed a weak humoral response after three doses of the mRNA-1273 vaccine and received a fourth dose. None of these patients had a previous history of COVID-19. The median interval from transplantation to the fourth dose was 6.1 years (IQR: 2.2−11.4 years). As for immunosuppressive therapy, 97% of the study patients were being treated with calcineurin inhibitors, 82% with mycophenolate mofetil, 76% with steroids, and 18% with mTOR inhibitors (Table 1). The median interval between the third and the fourth doses was 68 days (IQR: 63−82 days). After the fourth dose, the anti-RBD median titer increased significantly (p<0.0001) from 2.6 BAUs/mL (IQR: 13−66.3 BAUs/mL) to 112.5 BAUs/mL (IQR: 13.5−260 BAUs/mL). In parallel, median IC50 titers increased significantly (p=0.0001) from <7.5 (IQR : <7.5−15.1) to 47.1 (IQR <7.5−284.2). While only 16% of patients (n = 11) harbored neutralizing antibodies against the delta strain prior to the fourth injection, this percentage raised to 66% (n = 44) afterwards (Figure 1). Patients with undetectable neutralizing antibodies after the third dose were less likely to have their neutralizing capacity increased after the fourth dose (median fold-change: 1.1 *versus* 6.3, p=0.01). Notably, patients under combined therapy with tacrolimus, mycophenolate mofetil, and steroids displayed lower NAbs titers (median IC50 titers: 25.6 *versus* 140.6, p=0.01). Interestingly, NAbs titers against the delta variant were positively correlated with anti-RBD titers (Pearson’s r^2^=0.54, p<0.0001, Figure 2). Once the anti-RBD titer exceeded 143 BAUs/mL after the fourth dose, the specificity, sensitivity, positive predictive value, and negative predictive value to detect NAbs were 92.9%, 74.3%, 93.6%, and 72.2% respectively. No major adverse events were observed after the fourth dose. One patient who lacked neutralizing activity after the fourth injection (median IC50 titer: <7.5) developed symptomatic COVID-19 caused by the delta variant.

**Table 1.** General characteristics of the study patients according to the NAbs response after the fourth dose

|  | Entire cohort (n=67) | NAbs titer below 30<br>(n=28) | NAbs titer above 30<br>(n=39) | p |
| --- | --- | --- | --- | --- |
| Age (years) | 56.6 (47–64.6) | 53.1 (45–64.6) | 57.6 (50.6–64.6) | 0.29 |
| Male sex, n (%) | 41 (61.2%) | 12 (42.9%) | 29 (74.4%) | 0.01 |
| BMI (kg/m<sup>2</sup>) | 25.6 (22.3–29.6) | 24.3 (22.2–28.9) | 26.6 (22.7–29) | 0.48 |
| Comorbidities, n (%) |  |  |  |  |
| Cardiovascular disease | 19 (28.4%) | 8 (28.5%) | 11 (28.2%) | 0.59 |
| Diabetes | 27 (39.3%) | 10 (35.7%) | 17 (43.6%) | 0.61 |
| Hypertension | 53 (79.1%) | 21 (75%) | 32 (82%) | 0.56 |
| Time from kidney transplantation (years) | 6.1 (2.2–11.4) | 5.1 (1.8–11.1) | 7.5 (2.5–11.3) | 0.32 |
| First transplantation, n (%) | 54 (80.6%) | 23 (82.1%) | 31 (79.5%) | 1 |
| Deceased donor, n (%) | 55 (82.1%) | 22 (78.6%) | 33 (84.6%) | 0.54 |
| CNI, n (%) |  |  |  | 0.2 |
| Tacrolimus | 51 (76.1%) | 24 (85.7%) | 27 (69.2%) |  |
| Ciclosporin, n (%) | 14 (20.9%) | 3 (10.7%) | 11 (28.2%) |  |
| No CNI, n (%) | 2 (3%) | 1 (3.6%) | 1 (2.6%) |  |
| MMF/MPA, n (%) | 55 (82%) | 24 (85.7%) | 31 (79.5%) | 0.74 |
| mTOR inhibitors, n (%) | 12 (17.9%) | 4 (14.3%) | 8 (20.5%) | 0.74 |
| Steroids, n (%) | 51 (76.1%) | 24 (85.7%) | 27 (69.2%) | 0.15 |
| Tacrolimus +<br>MMF/MPA + steroids,<br>n (%) | 35 (52.2%) | 19 (67.9%) | 16 (41%) | 0.05 |
| Serum creatinine<br>(μmol/L) | 132 (106.2–162.5) | 131.3 (100.2-157.8) | 132 (115.3-162.6) | 0.61 |

Data are expressed as medians (interquartile ranges) unless otherwise indicated.
Abbreviations: BMI, body mass index; CNI, calcineurin inhibitors; MMF, mycophenolate mofetil; MPA, mycophenolic acid; mTOR, mammalian target of rapamycin; NAbs, neutralizing antibodies.

**Figure 1.**
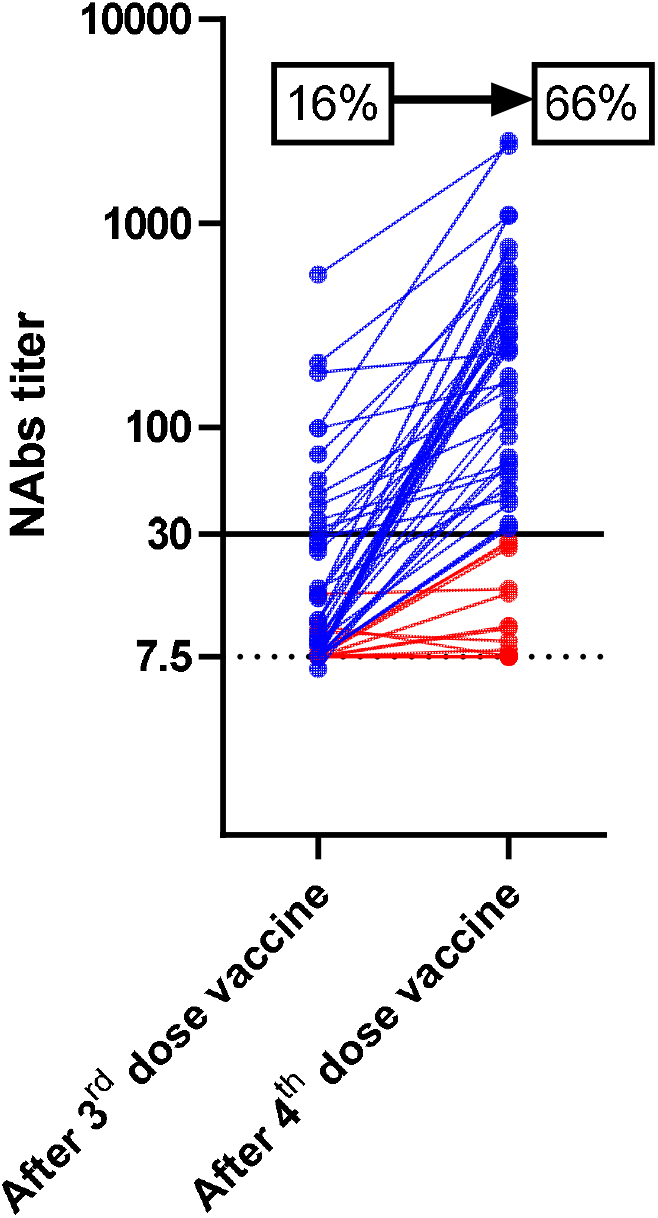
Neutralizing antibody titers against the delta variant after the third and fourth doses of the mRNA-1273 (Moderna) vaccine in 67 kidney transplant recipients. Neutralizing antibodies (NAbs) were expressed as the concentration capable of inhibiting 50% of the virus inoculum (EC50%; limit of detection: 7.5, dotted line). A titer >30 was considered as positive (black line). The percentages of patients harboring NAbs against delta variant before and after the fourth dose are shown in the graph. Lines in blue represent patients with NAbs with a titer is above 30 while red lines represent patients with NAbs with a titer is below 30

**Figure 2.**
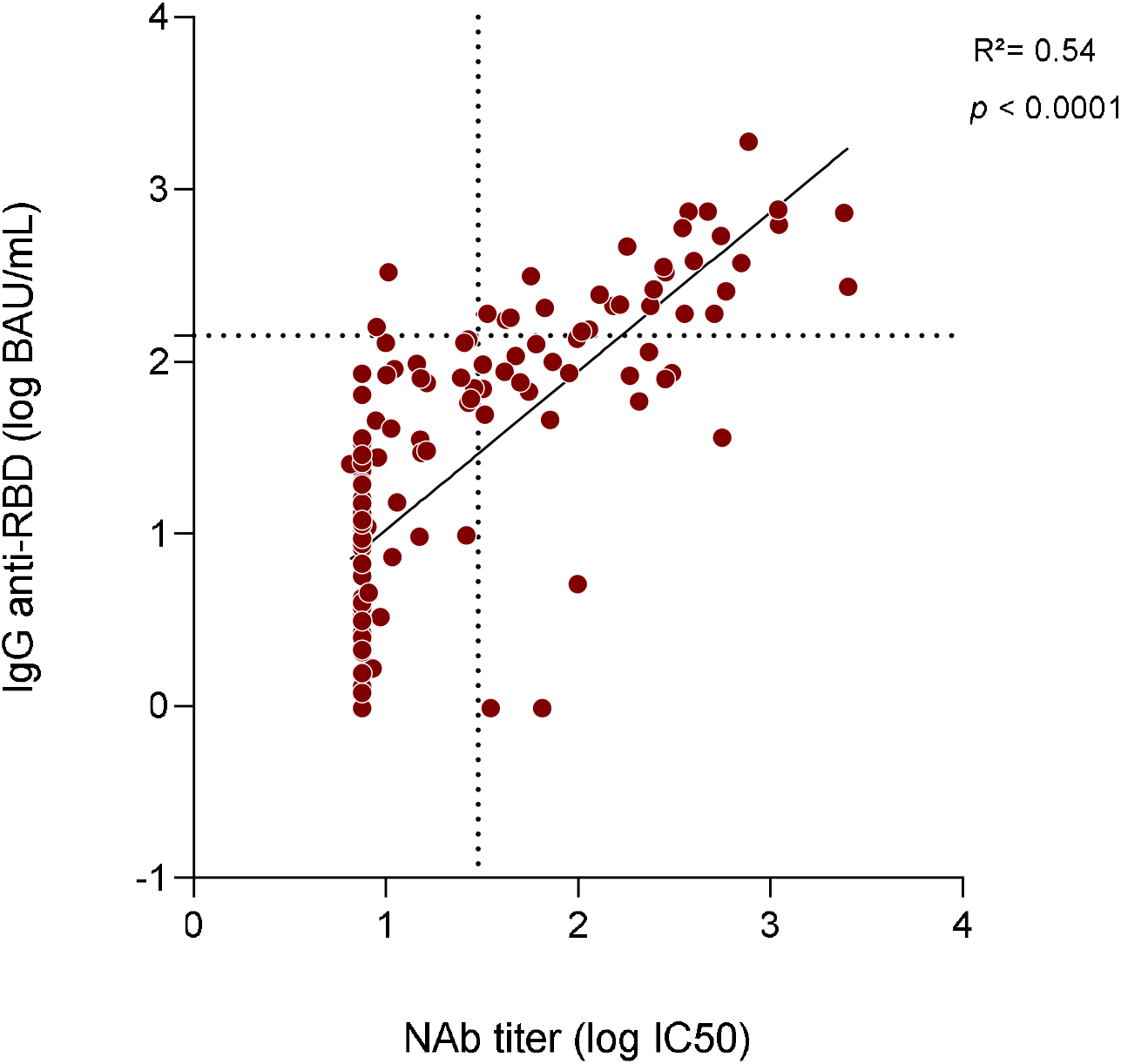
Scattergram and regression line showing a significant positive correlation between anti-receptor binding domain (RBD) IgG antibody titers (Log BAUs/mL) and neutralizing antibodies (NAbs) titers against the delta strain titer (log IC50) before and after the fourth dose of the mRNA-1273 (Moderna) vaccine in 67 kidney transplant recipients (Pearson r^2^=0.54, p < 0.0001). The dotted line parallel to the Y axis represents the cut-off for a positive NAbs titer (30), whereas the dotted line parallel to the X axis represents the cut-off for a weak RDB IgG antibody titer (143 BAUs/mL).

## Discussion

In this cohort of 67 kidney transplant recipients who showed a low immune response after three doses of the mRNA-1273 (Moderna) vaccine, a fourth dose significantly increased the neutralizing response against the currently dominant delta variant. Specifically, the proportion of patients harboring neutralizing antibodies against the delta strain rose significantly from 16% to 66%. Only one published study has focused on the effect of a fourth vaccine dose in a small cohort of 18 solid organ transplant recipients.^7^ In accordance with our data, the authors found that eight patients with negative or low response after the third dose developed high antibody titers after the fourth injection.^7^ Additionally, six of the ten patients who already had high positive titers after three doses showed further boosting after the fourth. However, this report did not provide data on the neutralizing activity – which has been shown to be highly predictive of immune protection against symptomatic SARS-CoV-2 infection.^8^

This is, to our knowledge, the first study to assess the effect of a fourth dose vaccine by taking into account the neutralizing activity against the delta variant. Previous research has focused on the response of solid organ transplant recipients after a third vaccine dose.^1,2,9,10^ While improvements have been observed for both B and T responses, a significant proportion of non-responders (30–50%) was reported. In a double-blind, randomized, placebo-controlled trial of a third dose of mRNA-1273 vaccine, Hall et al.^3^ found a higher neutralizing activity in the vaccine group *versus* the placebo group (median percent virus neutralization: 71% *versus* 25%, respectively). These observations suggest the existence of “an antigen dose effect” and that stronger immune stimuli are needed to improve vaccine immunogenicity. There have been previous reports of better immunogenicity after additional vaccines doses or higher vaccines doses in immunocompromised patients.^11,12^ Although a consensus threshold for COVID-19 seroprotection has not been defined yet, higher antibody titers have been correlated with a reduced risk of symptomatic infection.^13,14^ These data, coupled with our findings, indicate that a fourth vaccine injection is beneficial in transplant recipients who show a weak response after three doses. It is nonetheless concerning that a fourth injection was still unable to elicit neutralizing antibodies against this variant in approximately one third of our study sample. These patients should maintain strict sanitary protection measures and/or be considered as candidates for prophylactic administration of monoclonal anti-SARS-CoV-2 antibodies. While immunosuppressive modulation during vaccination could be another alternative to improve vaccine response, this approach should be weighed against the risk of developing acute rejection. No data are currently available on the cost-effectiveness of this strategy.

In summary, our data indicate that a fourth mRNA-1273 vaccine injection in kidney transplant recipients with a weak antibody response after three previous doses improves serum neutralization against the delta variant.

## Methods

The serological response after the third vaccine dose was monitored in our entire kidney transplant cohort. Patients with a weak serological response one month after the third dose were eligible for a fourth injection. Anti-IgG antibody titers were measured with an ARCHITECT IgG II Quant test (Abbott, Abbott Park, IL, USA; assay range: 1–11360 binding antibody units [BAUs]/mL). In light of previous data showing that IgG titers >143 BAUs/mL were associated with the presence of neutralizing antibodies against the wild-type virus and its common variants (alpha, beta, and gamma),^15^ titers <143 BAUs/mL were considered to reflect a weak humoral response. Neutralization of live viruses (delta strain) was investigated using S-Fuse reporter cells as previously described.^5^ Neutralizing antibodies (NAbs) against the delta strain were expressed as the concentration capable of inhibiting 50% of the virus inoculum (EC50%; limit of detection: 7.5). A titer >30 was considered as positive. Anti-RBD IgG and NAbs were quantified one month after the third and fourth vaccine doses, respectively. Continuous data are presented as medians and IQR and analyzed using the nonparametric Mann-Whitney *U* or Kurskal Wallis tests (for unmatched pairs) and the Wilcoxon signed rank tests (for matched pairs). Categorical variables are expressed as counts and percentages and compared using the Fisher’s exact test. The Pearson’s correlation coefficient (ρ) was used to express the correlation between NABs and IgG levels against the receptor binding domain (RBD). All calculations were performed using GraphPad Prism, version 8.0 (GraphPad Inc., San Diego, CA, USA). A two-sided P value□<□.05 was considered statistically significant. The study protocol was approved by the local Ethics Committee (comité de protection des personnes de Strasbourg, France, identifier: DC-2013– 1990) and written informed consent was obtained from participants.

## Data Availability

All data produced in the present study are available upon reasonable request to the authors

## Author Contributions

Concept and design: Benotmane, Caillard.

Experiments: Bruel, Planas, Schwartz

Acquisition, analysis, or interpretation of data: All authors.

Drafting of the manuscript: Benotmane, Caillard.

Critical revision of the manuscript for important intellectual content: All authors.

Statistical analysis: Benotmane

Administrative, technical, or material support: All authors

Supervision: Caillard.

